# Sixteen years of innovation in youth mental healthcare in Australia: Outcomes for young people attending headspace centre services

**DOI:** 10.1101/2022.08.24.22279102

**Authors:** Debra Rickwood, Juliet McEachran, Anna Saw, Nic Telford, Jason Trethowan, Patrick McGorry

## Abstract

Australia’s headspace initiative is world-leading in nation-wide youth mental healthcare reform for young people aged 12 to 25 years, now with 16 years of implementation. This paper examines changes in the key outcomes of psychological distress, psychosocial functioning, and quality of life for young people accessing headspace centres across Australia for mental health problems. The design was a single-arm, observational study using routinely collected data from headspace clients commencing an episode of care between 1 April 2019 and 30 March 2020, and at 90-day follow-up. All 108 of the fully established headspace centres across Australia were included, with 58,233 young people aged 12-25 years first accessing headspace centres for mental health problems during the data collection period. Main outcome measures were self-reported psychological distress and quality of life, and clinician-reported social and occupational functioning. Most headspace mental health clients presented with depression and anxiety issues (75.21%). There were 35.27% with a diagnosis: 21.74% diagnosed with anxiety, 18.51% with depression, and 8.60% were sub-syndromal. Cognitive behavioural therapy was the most common treatment. There were significant improvements in all outcome scores over time (P<0.001). From presentation to last service rating, over one-third of young people had significant improvements in psychological distress and a similar proportion in psychosocial functioning; just under half improved in self-reported quality of life. Significant improvement on any of the three outcomes was shown for 70.96% of headspace mental health clients. After 16 years of headspace implementation, positive outcomes are being achieved, particularly when multi-dimensional outcomes are considered. A suite of outcomes that capture meaningful change for young people’s quality of life, distress and functioning, is critical for early intervention, primary care settings with diverse client presentations, like the headspace youth mental healthcare initiative.

## Introduction

The mental health of young people in their teenage and early adult years is a major concern world-wide, due to the high and increasing prevalence of mental health problems at this time of life [1]. Concern has escalated in the context of the disproportionate impact of COVID-19 on this demographic group through major disruptions to their studies, work opportunities, and social relationships [2]. An international movement for youth mental health services, tailored specifically to the needs of the 12-25-year age range, emerged around 20 years ago and has expanded rapidly [3]. Australia was one of the first countries to invest in national reform by creating a new health service platform for young people through headspace: the National Youth Mental Health Foundation [4]. headspace now comprises the largest national network of enhanced primary care, youth mental health services, world-wide, with over 135 headspace centres across Australia [5]. Similar service approaches in youth mental healthcare are being implemented in many other countries, including Ireland, Canada, Denmark, Israel, the United Kingdom, and parts of the United States [6].

headspace commenced in 2006, funded by the Australian Government, with the first 10 centres established in 2007. Centres have continued to be implemented over the past 16 years, and there are currently 149 centres open across Australia (headspace communication 6 April 2022). This upscaling has been in response to strong support from communities and vocal advocacy to have centres established in local areas, as well as bi-partisan support from the Australian Government. The headspace centre network has been further augmented by the introduction of online services in 2011 (eheadspace), early psychosis services in some locations from 2014, online work and study support since 2016 and, more recently, centre service innovations such as satellites and remote outreach. All of these different ways to expand capacity in youth mental healthcare have been necessary to reach and address the varied needs of young people in diverse communities across the wide expanse of Australia [7].

The foundation of the headspace initiative is the headspace centre network. headspace centres have been co-designed with young people to break down the barriers that young people typically experience to accessing in-person mental healthcare, including lack of mental health literacy and uncertainty regarding need, stigma, fears about confidentiality, cost, and poor experiences of care [4, 8]. Centres provide easy-access, youth-friendly, integrated primary care services, with four core streams of service delivery to holistically address the main health and wellbeing needs for young people aged 12-25 years—mental health, physical and sexual health, alcohol and other drugs, and work and study issues. The centre model was designed primarily for young people with mild to moderate common mental health problems, and to encourage them to seek help early in the development of problems. Notwithstanding, centres have a #x2018;no wrong door’ approach so that young people are supported to access support as early and easily as possible whatever their mental health status. This recognises that young people at all stages of illness experience significant access barriers to mental healthcare. Importantly, young people can self-refer or be referred by other services or by family and friends.

The headspace centre model comprises 16 core components, including 10 service components (e.g., youth participation, family and friends participation, community awareness, early intervention, supported transitions) and six enabling components (e.g., blended funding, monitoring and evaluation, national network), which are more fully described elsewhere see [9]. Briefly, the key innovations of the model, embraced by the youth mental healthcare movement internationally, include the focus on the 12-25-year age range, rather than the traditional child and adolescent (under 18 years) versus adult (18-65 years) demarcation for mental health services. Youth participation and engagement are prioritised at all levels, including young people driving decisions about their own care; youth input to service design, delivery and evaluation; and the embedded involvement of young people in governance processes. The participation of family and friends is also prioritised and actively supported. Centres must bring together access to the four core streams of service delivery in one location, and within a primary care focus, although strong integration with secondary and tertiary services is also required. Youth-friendliness/focus is recognised as critical. This is achieved through young people’s input during establishment and maintenance of the service; by ensuring that the look and feel of the centre is welcoming, positive and engaging for young people; that young people feel accepted and respected for who they are; and that the service is designed around the needs of young people in the local community. Two additional priorities are particularly unusual for mental healthcare. These are a requirement for community awareness activities, which are specifically funded to improve mental health literacy and pathways to care for young people in the local community; and brand adherence, which is strongly promoted and carefully maintained to ensure fidelity and credibility.

The effectiveness of headspace and the impact of the Australian Government’s investment into the initiative are of considerable interest in Australia and world-wide. headspace centres have undergone three external evaluations funded by government. A preliminary external evaluation in 2009 of the first 30 centres showed that young people found the approach to be acceptable [10]. A 2015 evaluation reported that centres: were highly accessible and utilised by a diverse range of young people with high psychological distress; facilitated access for young people living outside major cities; demonstrated a statistically significant small program effect; and that young people whose mental health improved also had positive economic and social outcomes and reduced suicidal ideation [11]. A third government-commissioned external evaluation will be completed in 2022, and preliminary drafts show continued strong youth and community support.

Importantly, none of these well-resourced external evaluations have been able to identify an appropriate control or comparison group. Given that headspace is a government-funded, real-world, service system innovation, which is delivering currently available evidence-based mental healthcare, and being rapidly scaled up to meet the high level of need and demand across Australia, randomised trials are neither feasible nor appropriate. Comparative data from young people accessing no care or other types of care are very difficult to attain or are not appropriate comparators. The closest comparator in Australia has been the Access to Allied Psychological Services (ATAPS) initiative, also government funded. A comparative study, using routinely collected data from 2009-2012, concluded that ATAPS and headspace both delivered free or low-cost psychological services to 12-25 year-olds with different characteristics, and that both had promising effects on mental health, filling a service gap for young people in a complementary way [12]. Into the furture, however, the best comparisons will be with findings from other similar services being implemented internationally.

Notably, headspace has prioritised internal evaluation from the outset, introducing an innovative routine data collection system in 2013 to gather information from all young people accessing centres and their service providers at each occasion of service, and attempting to follow-up young people 90 days after service exit. These data have been used to describe the characteristics and presenting issues of young people accessing centres[13], the services provided to them [14], as well as an early report of key outcomes [15].

Determining appropriate outcomes, which can be collected routinely for a large cohort of young people with very diverse demographic characteristics, clinical presentations, and service needs, is a challenging task. A systematic review of literature published up until mid-2014 revealed no mental health outcome measures designed specifically for the 12-25-year age range [16]. The review also highlighted the many different facets of mental health that could be the focus of outcome measurement, including measures of disorder-specific symptoms, global cognition and emotion measures, functioning, quality of life, recovery, as well as multidimensional constructs.

headspace initially implemented, as key outcome measures, a self-reported measure of psychological distress, the Kessler 10 Psychological Distress Scale (K10 [17]), and a clinician-reported measure of social and occupational functioning, the Social and Occupational Functioning Assessment Scale (SOFAS [18]). These were included based on the requirements for government funding, and widespread consultation with the headspace network to determine brief, global assessments of mental health status relevant to young people.

An initial outcomes analysis was published in 2015 [15] based on these two measures. In that study, we analysed the K10 and SOFAS scores for 24,034 young people commencing an episode of care between 1 April 2013 and 31 March 2014 for mental health problems at one of the 55 then-established centres [15]. Results showed that over one-third of young people had significant improvements in psychological distress (K10) and a similar proportion improved in psychosocial functioning (SOFAS); 60% showed significant improvement on either measure.

Subsequently, to fill the gap of no young-person centric, global, mental health outcome measure, headspace co-developed with headspace young people and service providers, a measure called MyLifeTracker (MLT) [19]. This is a brief self-report measure of quality of life in five domains of importance for young people, which can be used for routine outcome monitoring and measurement informed treatment, as well as for tracking outcomes at the service level [20]. Although still not sufficiently comprehensive to capture the diverse nature of early intervention outcomes for youth mental heathcare, the three measures of self-reported quality of life (MLT), self-reported psychological distress (K10), and clinician-reported social and occupational functioning (SOFAS), altogether, comprise a suite of outcome measures appropriate for routine collection within headspace centre services that provide information about change in the relevant, multiple dimensions of distress, quality of life, and functioning.

The headspace centre network is now 16 years old, with over 700,000 young Australians having received services since its inception [5]. The current study aims to describe multi-dimensional outcomes for young people accessing mental healthcare at headspace centres in terms of changes in their psychological distress, quality of life, and social and occupational functioning. We examine significant change, reliable change, and clinically significant change scores, as increasingly conditional indicators of change, for the three outcomes. Describing the nature of the outcomes being achieved by headspace centre services in Australia contributes to the growing international literature in youth mental healthcare models and provides comparative data for similar services internationally.

## Method

### Participants

Participants were all young people who commenced and completed an episode of care at a headspace centre for mental health reasons between 1 April 2019 and 30 March 2020. We selected this time period to avoid the impact of COVID-19 on both clients, themselves, and the routine program data collection. Like many services, headspace was severely impacted by COVID-19 restrictions, including having to deliver services exclusively online and via phone in many areas for considerable periods of time. This caused difficulties collecting data from young people in the changed service delivery circumstances.

Only young people who attended mental health services were included. Young people can also attend headspace for situational, physical or sexual health, alcohol or other drug, and vocational reasons without needing to access mental health care, so young people with these reasons for presentation were excluded (n=18631). Figure 1 shows the sample numbers at each point of the data selection process.

**Fig 1.** Number of clients by each data selection process.

This yielded an overall sample of 58233 clients across the 108 headspace centres that were fully operational during the data collection period. The majority of clients were female (61.34%), 36.96% were male, and 1.70% were intersex or transgender. Average age was 17.43 years (SD=3.48), with 24.27% aged 12-14, 30.07% aged 15-17, 23.32% aged 18-20, and 22.33% aged 21-25 years.

### Procedure

The design was a single-arm, observational study using routinely collected service data. headspace collects a minimum data set (MDS) from young people and their service providers at every occasion of service [21]. Young people are asked to complete a series of questions while waiting for their service visit; this is usually done on a tablet device or at a stand-alone computer located in part of the reception area that gives privacy for data entry. Young people are directed to the data collection process by reception staff, unless they require support to enter their information, in which case they are assisted by a service provider if they agree to this. Service providers enter data about each occasion of service into the MDS system after each service.

Follow-up data on psychological distress are collected after a 90-day break in service provision from young people who volunteer when they first attend headspace to participate in a follow-up questionnaire. Those who agree are emailed or texted a follow-up questionnaire to self-complete. There were 4.08% (n=1574) of eligible young people who responded to the follow-up survey.

All the routinely collected MDS data from every headspace centre are stored in secure data warehouse, and anonymous data for each client at each occasion of service are available to headspace National research and evaluation staff for analysis.

Ethical approval was obtained from Melbourne Health Quality Assurance Review and the headspace Data Governance Reference Group.

### Measures

Demographic characteristics of gender and age were self-reported or completed by clinicians at first visit. Gender options included: male, female, and gender diverse (comprising 8 categories). Age was determined by date of birth and categorised into early adolescence (12-14 years), mid-adolescence (15-17 years), late adolescence (18-20 years), and early adulthood (21-25 years).

Clinical characteristics of the clients were determined by clinicians at each visit, and included: primary presenting issue, indicated on a list of 13 mental health and behaviour concerns (including an open-ended ‘other’ category); primary and other diagnosis based on DSM-V categories; mental health risk status, indicating whether the young person had ‘No risk factors or symptoms of mental health problems’, ‘Risk factors present’, ‘Sub-threshold symptoms’, ‘Threshold diagnosis - first episode’, or ‘Threshold diagnosis - ongoing mental disorder’; and type of treatment, determined via a list of 50 treatment types.

Client outcomes were: psychological distress, self-reported through the 10-item Kessler Psychological Distress Scale (K10)[17]; overall psychosocial functioning, assessed by service providers using the single-item Social and Occupational Functioning Assessment Scale (SOFAS)[18]; and a self-reported quality of life measure developed specifically for youth mental health services, the 5-item MyLifeTracker (MLT)[19]. Psychological distress is self-reported immediately before young people’s first to fourth, seventh, eleventh, and fifteenth visits, and at 90-day follow-up; quality of life is self-reported at every visit; and psychosocial functioning is recorded by service providers at every visit.

### Statistical Analyses

Significance was set at P<0.001 to exclude trivial effects being significant due to high statistical power.

Changes in outcomes were assessed in two ways [22]. First, mixed-design analysis of variance (ANOVA) determined change over time in K10, SOFAS and MLT scores by time (first/last assessment), number of sessions, age-group (early/mid/late adolescence and early adulthood) and gender (male/female; the gender diverse group was not included in analyses due to small sample sizes in the groups). Associations between outcome scores were determined with Pearson’s correlation coefficient. Logistic regression examined differences between those who did and did not achieve significant improvement on K10, MLT or SOFAS and those who did or did not provide follow-up data.

Second, significant change, reliable change, and clinically significant change scores were calculated for each of the three outcomes, as increasingly conditional indicators of change. Significant change was determined by a change with at least a moderate effect size (0.5). The Jacobson and Truax method [23] was used to determine reliable change (RCI) (indicating reliable improvement or decline) and clinically significant change (CSI) (cut-off point at which the young person is more likely to belong to a non-clinical than clinical population). The RCI and CSI for the K10 and SOFAS were calculated as for the 2015 analysis: RCI=7 and 10, CSI=23 and 69, respectively [15]. For MLT, these were RCI=18.27, CSI=63.86 [19, 20].

## Results

### Clinical characteristics and treatment services

The most common primary presenting issues were anxiety symptoms (41.85%) followed by depression symptoms (33.36%), together accounting for three-quarters of primary presenting concerns (see Table 1). This was evident for all age and gender groups, except 12-14 year-old boys, whose second most common presenting issue was anger issues (23.36%).

**Table 1.** Percentage of young people with each primary presenting issue by gender and age-group.

| Age | n | Anxiety symptoms | Depression symptoms | Anger Issues | Stress related | Suicidal thoughts/behaviour | disruptive behavioural issues | Difficulties with personal relationships | Conflict in home environment | Borderline personality traits | Eating disorder |
| --- | --- | --- | --- | --- | --- | --- | --- | --- | --- | --- | --- |
| Female |  |  |  |  |  |  |  |  |  |  |  |
| 12-14 | 7062 | 46.12 | 26.90 | 8.61 | 5.44 | 3.40 | 2.11 | 2.25 | 3.41 | 0.34 | 1.36 |
| 15-17 | 9321 | 44.50 | 34.32 | 3.75 | 6.00 | 2.84 | 0.75 | 1.87 | 2.42 | 1.52 | 2.02 |
| 18-20 | 7167 | 45.22 | 34.70 | 2.05 | 6.60 | 2.51 | n/a | 1.88 | 1.16 | 3.38 | 2.13 |
| 21-25 | 6307 | 44.47 | 34.75 | 2.24 | 7.15 | 1.81 | n/a | 2.51 | 0.73 | 3.95 | 2.01 |
| Male |  |  |  |  |  |  |  |  |  |  |  |
| 12-14 | 4221 | 34.66 | 21.37 | 23.36 | 5.33 | 2.11 | 8.22 | 1.99 | 3.10 | 0.07 | n/a |
| 15-17 | 5089 | 34.60 | 35.53 | 12.7 | 6.92 | 3.30 | 2.97 | 1.69 | 1.89 | n/a | n/a |
| 18-20 | 4067 | 39.32 | 39.56 | 6.96 | 5.88 | 3.49 | 1.08 | 1.89 | 0.64 | 0.79 | n/a |
| 21-25 | 4213 | 37.43 | 41.11 | 6.74 | 6.34 | 2.71 | 0.88 | 2.60 | 0.71 | 1.09 | n/a |
| Total |  |  |  |  |  |  |  |  |  |  |  |
| 12-25 | 47447 | 41.85 | 33.36 | 7.26 | 6.22 | 2.77 | 1.79 | 2.01 | 1.85 | 1.59 | 1.31 |

Notes: Total sample size differs from total possible sample size of 58223 due to missing data on gender and age, and the exclusion of 'gender diverse' due to small cell sizes. n/a cell size<30.

Mental health risk status (Table 2), which shows the highest level of risk recorded across the episode of care, revealed that almost a quarter (23.57%) had no indicated risk factors or symptoms of mental health problems; most of these young people attended only an initial consultation and did not progress to receiving treatment services. Another quarter (25.64%) had identified risk factors; one fifth (21.01%) had sub-threshold symptoms; 11.77% were first episode; and 18.01% had full-threshold diagnosis with ongoing mental disorder.

**Table 2.** Mental health risk status.

| Mental health risk status | Young people presenting with mental health and behavioural issues |  |
| --- | --- | --- |
|  | n | % |
| No risk factors or symptoms or mental health problems | 13724 | 23.57 |
| Risk factors present | 14928 | 25.64 |
| Sub-threshold symptoms | 12230 | 21.01 |
| Threshold diagnosis – first episode | 6856 | 11.77 |
| Threshold diagnosis – ongoing mental disorder | 10458 | 18.01 |
| **Total** | **58223** | **100.0** |

Consistent with risk status, Table 3 shows that no diagnosis was made for two-thirds of clients (64.71%), including 14.31% with diagnosis not yet assessed and 8.60% who were sub-syndromal. For those with a diagnosis, the most common was anxiety disorder (21.74%) followed by depressive disorder (18.51%), then trauma (5.64%), personality disorder (2.34%) and disruptive behaviour (2.03%). All other diagnoses were proportionally very rare.

**Table 3.** Any diagnosis during the episode of care.

| Diagnosis | Young people presenting with mental health and behavioural issues |  | Young people with at least 2 outcome measures for K10/MLT or SOFAS |  |
| --- | --- | --- | --- | --- |
|  | n | % | n | % |
| Missing | 6 | 0.01 | 2 | 0.0 |
| Not applicable (diagnosis not relevant or service provider not qualified to give diagnosis) | 23433 | 40.24 | 9615 | 37.0 |
| Diagnosis not yet assessed | 8332 | 14.31 | 4218 | 16.2 |
| No diagnosis and no sub-syndromal symptoms | 897 | 1.54 | 427 | 1.6 |
| No diagnosis but sub-syndromal mental health problems | 5008 | 8.60 | 2170 | 8.3 |
| **Total no diagnosis** | **37421** | **64.71** | **16432** | **63.2** |
| Anxiety | 12663 | 21.74 | 3993 | 15.4 |
| Depression | 10777 | 18.51 | 3398 | 13.1 |
| Trauma | 3284 | 5.64 | 918 | 3.5 |
| Personality disorder | 1365 | 2.34 | 337 | 1.3 |
| Disruptive behaviour | 1180 | 2.03 | 330 | 1.3 |
| Feeding disorder | 788 | 1.35 | 159 | 0.6 |
| Substance use disorder | 705 | 1.21 | 129 | 0.5 |
| Obsessive compulsive | 579 | 0.99 | 148 | 0.6 |
| Bipolar disorder | 389 | 0.67 | 114 | 0.4 |
| Schizophrenia | 317 | 0.54 | 62 | 0.2 |
| Somatic disorder | 113 | 0.19 | 15 | 0.1 |
| Other | 3373 | 5.79 | 1412 | 5.4 |
| **Total any diagnosis** | **20547\*** | **35.29** | **9578\*** | **36.8** |
| **Total Young People** | **58233** | **100** | **26010** | **100** |

Notes: \*Young people can have more than one primary diagnosis over their episode of care, so diagnoses do not sum to the total.

**Table 4.** Most common mental health treatment types.

| Treatment type | % of treatments | N of treatments |
| --- | --- | --- |
| Cognitive behavioural therapy | 31.23 | 70690 |
| General or supportive counselling | 12.53 | 28363 |
| Acceptance and commitment therapy | 5.47 | 12384 |
| Psychoeducation | 4.79 | 10842 |
| Skills training (social and communication skills anger management) | 3.32 | 7507 |
| Interpersonal therapy | 3.26 | 7385 |
| Motivational Interviewing/Enhancement | 2.24 | 5072 |
| Behavioural interventions (including activity scheduling exposure techniques) | 2.20 | 4980 |
| Cognitive interventions (eg Cognitive analytic therapy) | 1.65 | 3732 |
| Problem solving therapy | 1.34 | 3022 |
| **Total treatments<sup>2</sup> delivered** | NA<sup>1</sup> | **226307** |
| **Median number of treatments recorded per young person** |  | **2.00** |

Notes: <sup>1</sup>% do not sum to 100 because young people can receive more than one service type and only the 10 most common treatment types are listed. <sup>2</sup>Service providers can record up to three treatment types per visit.

The most common treatment was cognitive behaviour therapy, comprising 31.23% of treatments. General or supportive counselling was next (12.53%), and then acceptance and commitment therapy (5.47%).

### Mean changes in outcomes over time

The mean outcome scores at each time point (visit number) for young people who had received that number of sessions are plotted in Figures 2-4. The respective sample sizes show that the sample size declined dramatically with each time point.

**Fig 2.** Mean psychological distress (K10) scores by time point and number of sessions.

**Fig 3.** Mean social and occupational functioning (SOFAS) scores by time point and number of sessions.

**Fig 4.** Mean quality of life (MLT) scores by time point and number of sessions.

For the K10, the strongest effect was the main effect for time, explaining 7.51% of the variance. Other effects were significant but negligible (1.0% variance or less). When the 90-day follow-up was added, the time effect explained 12.43% of the variance. Note that while the follow-up sample was generally similar to the whole sample, they were more likely to be female, OR=2.00 (95% CI=1.73-2.23); and more likely to be older, OR=1.03 (95% CI=1.01-1.05).

For SOFAS scores, again the time effect was strongest, although only explained 2.76% of the variance; no other sizeable effects were evident.

For MLT, the time effect explained 11.73% of the variance, and age-group explained 2.08%.

### Significant, reliable, and clinically significant change

The percentage of young people showing significant, reliable, and clinically significant change from first to last rating are presented in Table 5. For psychological distress, just over one-third significantly, a quarter reliably and one-fifth clinically significantly improved, and 5.94% reliably deteriorated on the K10. For quality of life, MLT scores revealed just under half significantly improved and about 30% each showed reliable and clinically significant change; 5.03% had a reliable deterioration. According to clinician ratings, 36.12% significantly, 29.92% reliably and 37.81% clinically significantly improved in psychosocial functioning (SOFAS); 13.84% reliably declined.

**Table 5.** Percentage of young people showing significant, reliable, and clinically significant change in outcomes from first to last service rating.

| Outcome | Change method | N | Change Category |  |  |
| --- | --- | --- | --- | --- | --- |
|  |  |  | Improvement % | No change % | Deterioration % |
| **K10** | Significant Change | 22349 | 35.50 | 54.27 | 10.23 |
|  | Reliable Change | 22349 | 24.53 | 69.53 | 5.94 |
|  | Clinically Significant Change | 17236 | 22.39 | 77.61 | NA<sup>1</sup> |
| **SOFAS** | Significant Change | 24997 | 36.12 | 46.35 | 17.53 |
|  | Reliable Change | 24997 | 29.92 | 56.24 | 13.84 |
|  | Clinically Significant Change | 15501 | 37.81 | 62.19 | NA<sup>1</sup> |
| **MLT** | Significant Change | 22203 | 46.85 | 41.49 | 11.65 |
|  | Reliable Change | 22203 | 30.05 | 64.92 | 5.03 |
|  | Clinically Significant Change | 17549 | 29.87 | 70.13 | NA<sup>1</sup> |
| **For any outcome<sup>2</sup>** | Significant Change | 21053 | 70.96 | 13.18 | 31.47 |
|  | Reliable Change | 21053 | 56.35 | 29.39 | 21.49 |
|  | Clinically Significant Change | 10118 | 51.72 | 48.27 | NA<sup>1</sup> |

Notes: <sup>1</sup>Young people in the non-clinical population at first time (20.88% of 22349 for K10, 37.99% of 24997 for SOFAS, and 20.96% of the 22203 for MLT) are unable to make a clinically significant improvement and are therefore excluded. Young people in the clinical population are not able to deteriorate, but rather remain in the clinical population (so included in 'No change' category).
<sup>2</sup> For any of the three outcomes, the rows do not sum to 100% because young people can be in more than one category.

The K10, SOFAS and MLT measure different aspects of mental health; and psychological distress and quality of life are self-reported by young people, while social and occupational functioning is estimated by the clinician. The K10 and MLT are moderately strongly correlated at both first and last assessment: *r=*-0.65 and 0.64, respectively. These measures both correlate weakly with SOFAS scores at first (K10 *r=*-0.23; MLT *r=*-0.24) and last (K10 *r=*-0.29; MLT *r=*-0.34) assessment.

If all three measures are considered, 70.96% of young people significantly improved, 56.35% reliably improved, and 51.72% clinically significantly improved. There were 31.47% who significantly deteriorated on any outcome measure. A logistic regression, summarised in Table 6, comparing young people who improved with those who did not (significant improvement on at least one measure) showed that improvement was predicted by being male and attending a greater number of sessions (visits). Improvement was also predicted by greater distress (OR, 1.01: 95% CI, 1.01-1.02), lower psychosocial functioning (OR, 0.97: 95% CI, 0.96-0.97), and lower quality of life (OR, 0.99: 95% CI, 0.99-0.99) at baseline.

**Table 6.** Summary of logistic regression model predicting significant improvement on any outcome (K10, MLT or SOFAS).

| Factor |  | OR | CI | P |
| --- | --- | --- | --- | --- |
| Gender | Male | 1.13 | 1.05-1.20 | 0.001 |
|  | Female (1) |  |  |  |
| K10 baseline |  | 1.01 | 1.01-1.02 | <0.001 |
| MLT baseline |  | 0.99 | 0.99-0.99 | <0.001 |
| SOFAS baseline |  | 0.97 | 0.96-0.97 | <0.001 |
| Number of mental health visits |  | 1.15 | 1.13-1.16 | <0.001 |
| Age | 12-14 years | 0.97 | 0.88-1.06 | 0.46 |
|  | 15-17 years | 0.90 | 0.82-0.98 | 0.02 |
|  | 18-20 years | 0.95 | 0.87-1.05 | 0.32 |
|  | 21-25 years (1) |  |  |  |
| Primary presenting concern: |  |  |  |  |
| Anxiety |  | 1.05 | 0.94-1.16 | 0.39 |
| Depression |  | 1.01 | 0.91-1.13 | 0.81 |
| Anger |  | 1.11 | 0.96-1.30 | 0.17 |
| Stress |  | 1.04 | 0.89-1.21 | 0.66 |
| Suicidal |  | 1.10 | 0.85-1.41 | 0.48 |
| Difficulty in personal relationships |  | 1.20 | 0.97-1.49 | 0.09 |
| Attention deficit/behavioural |  | 1.12 | 0.84-1.48 | 0.45 |
| Conflict |  | 0.98 | 0.79-1.21 | 0.83 |
| Borderline personality traits |  | 0.71 | 0.52-0.99 | 0.04 |
| Eating disorder |  | 0.71 | 0.52-0.97 | 0.03 |

Notes: (1) Indicates reference group. Overall $X^2 [5] = 1473.23$ , Nagelkerke R square=0.098

## Discussion

Our aim was to examine outcomes for headspace centre clients after 16 years of implementation and expansion of the network to 108 services, thereby making the outcomes attained transparent to the Australian community and providing comparative data for similar international initiatives. The results are generally comparable to the client outcomes reported in 2015 [15], although the number of centres has more than doubled since then along with the number of clients receiving services within the 12-month analysis period.

Interestingly, anxiety symptoms were the most common primary presenting issue in 2019-2020 followed by depressive symptoms, whereas the opposite order was evident in 2015. The high prevalence of anger issues for 12-14 year-old boys has remained as their second most common main presenting issue. This shows a need to focus on this particular form of mental distress for the youngest boys accessing headspace. Other headspace research confirms that early adolescent boys appear to be quite a distinct group with unique needs [20].

We reveal that only about one-third of headspace centre clients had a diagnosis, but 25.64% had risk factors present and 21.01% had sub-syndromal mental health conditions, reflecting the early intervention focus of headspace centres, which aim to intervene before symptoms have reached diagnostic thresholds. This also likely reflects that headspace service providers are primarily allied health workers, mostly psychologists, many of whom work within a non-diagnostic practice framework. Primary diagnoses were consistent with presenting issues, showing a preponderance of anxiety and depression. Many other diagnoses were evident, however, albeit with much lower frequency, showing the breadth of presentations that headspace centres must be equipped to address.

This breadth of presentations, in terms of issues and severity, necessitates multidimensional outcome measures to be used. The inclusion in this study of MLT as a quality of life measure is especially important considering that different domains of young people’s lives can be both influenced by and determinants of their mental health status. Our results show that 70.96% of headspace clients improved on at least one of the three outcome measures. The quality of life measure shows the greatest proportion improving (46.85%) supporting the promotion of headspace centres as easy access facilities for any type of problem, and the value of a broad patient-reported outcome measure tapping domains of life that are of concern for young people—specifically, their overall wellbeing, relationships with friends and family, day-to-day functioning in work/study/leisure activities, and general coping with life capacity.

By necessity, the current study used a single-arm, observational design; consequently, a limitation is the lack of a control group. An equivalent control group has not been able to be determined by any of the external evaluations. Comparative data, in general, are difficult to find, because headspace clients present for a wide range of reasons, treatments are varied, and centres are uniquely adapted to their varied communities and circumstances. Consequently, few other services are comparable. Recent systematic reviews and meta-analyses of psychological treatments for depression in children and adolescents report that 60% of youth receiving therapy do not respond within two months, and that 39% respond in treatment conditions and 24% in control conditions [24]. For adults, almost 60% do not respond to treatment, 41% respond within two months, and 17% respond in control conditions [25]. These findings come from rigorous randomised controlled trials and only consider psychological treatments for depression, so do not provide direct comparators for headspace; but, given that one of the main presenting issues for headspace clients is depression, they do provide a reference point. The reviews also support the contention that the effects of psychotherapies for depression are smaller in children and adolescents than in adults.

Recently, information has begun to be reported internationally for other primary care, youth mental health initiatives similar to headspace. For example, the evolution of Jigsaw in Ireland [26] and early learnings from the Foundry in Canada [27] have been described. No outcome data are yet available, although a study protocol for the Canadian Access Open Minds youth services has been published [28].

Another limitation is that our results are likely to underestimate treatment effects because the outcome data are collected at the last recorded data point, which for some young people is not at the completion of their treatment. Further, data are collected as the client presents for their treatment session rather than at the end of the session.

Our follow-up rate is unsurprisingly disappointing, although the results suggest that young people continue to improve after leaving the service. It is a major challenge to get young people to complete follow-up information after they have left a service, particularly through routine data collection (as opposed to a well-resourced research project). Nevertheless, there was a large number of young people who provided 90-day follow-up data, if only a very small percentage. The follow-up sample is skewed towards those who are female, but does show maintenance of outcomes for reduced psychological distress. Unfortunately, MLT was not available at follow-up for these analyses, but has more recently been included in the follow-up data protocol.

The impact of COVID-19 has had many unfortunate impacts on most people, including headspace young people and particularly on data collection. Most headspace centre services had to rapidly change to online delivery after March 2020, and many centres had long periods where no in-person services were possible due to lockdowns. This had a major impact on data collection, which was previously undertaken while the young person was waiting in-person for their occasion of service. Online service delivery meant online data collection, and although centres were able to adapt to incorporate this, it has proved more challenging for data compliance. Hence, the data reported here are for the period immediately prior to the impacts of COVID-19.

headspace is committed to examining and reporting outcomes for young people accessing its services. We will continue to use our MDS to contribute to the well-acknowledged complex challenge of what works for young people, particularly for the common presentations of anxiety and depression [29], which are the main issues for young people coming to headspace. These latest centre results show a positive impact, particularly when considering multiple outcome measures, which we argue is needed to match the holistic early intervention intent of headspace services. In particular, MLT measures outcomes directly relevant to young people’s lives, and it is validating that these items demonstrate the strongest improvement.

Nevertheless, there remain many young people for whom the services currently provided by headspace are not sufficient, and further research is required to identify how the outcomes for these young people can be improved. In particular, young people who have been referred to as the ‘missing middle’ [30] present to headspace centres as a result of their easy access, ‘no-wrong-door’ approach. These young people are likely to be too complex and their issues too severe for the headspace indicated prevention focus; headspace centres were originally intended for mild to moderate presentations of high prevalence conditions. Due to the lack of appropriate services in most communities and the high visibility and youth-friendliness of headspace centres, young people with more challenging presentations than first anticipated are attending. Centres increasingly must hold young people with more acute, complex and enduring mental health problems, despite not being designed nor resourced to do so. Providing services to these young people may attenuate the outcomes achieved. Consequently, identifying and analysing outcomes for more complex headspace clients is a future research need.

In conclusion, the current study shows that Australia’s headspace initiative, which has been at the forefront of youth mental health service reform and provides an exemplar internationally, is achieving important outcomes for the majority of young people who attend, providing a much-needed reoriented service response to address the high and rising prevalence of mental health problems for young people in their adolescent and early adult years.

## Data Availability

All data produced in the present work are contained in the manuscript

## Acknowledgements

None.

## Supporting information

S1_Fig Fig 1. Number of clients by each data selection process.

S2_Fig Fig 2. Mean psychological distress (K10) scores by time point and number of sessions.

S3_Fig Fig 3. Mean social and occupational functioning (SOFAS) scores by time point and number of sessions.

S4_Fig Fig 4. Mean quality of life (MLT) scores by time point and number of sessions.

## Notes

### Competing Interest Statement

The authors have declared no competing interest.

### Funding Statement

This study did not receive any funding

### Author Declarations

Royal Melbourne Hospital Human Research Ethics Committee of the Royal Melbourne Hospital (Australia) gave ethical approval for this work.

